# Occupational dermatoses in health care personnel using PPE during the COVID pandemic

**DOI:** 10.1101/2023.07.13.23292421

**Authors:** Dinesh P Asati, Kapil Baheti, Maninder Kaur, Suman Patra, Kritika Singhal

**Author notes:** **Corresponding Author** Dr. Kapil Baheti, Junior Resident, Department of Dermatology, Venereology, and Leprosy, All India Institute of Medical Sciences, Bhopal. **Author approval** All the authors have seen and approved the manuscript. **Competing interests** There are no competing interests. **Funding statement** This study had no source of funding.

## Abstract

**Background:** A sudden surge of occupation-associated dermatoses among the healthcare workers (HCWs) serving COVID-19 patients have been witnessed recently due to increased usage of PPE (PPE) kits and increased frequency of hygiene practices, with a significant impact on their quality of life and compromised efficacy at work. Hence, this study was conducted to measure the prevalence of occupational dermatoses among HCWs serving Covid-19 patients using PPE kits and hygiene practices and their impact on quality of life.

**Methods:** HCWs of all cadres were screened for occupation-associated dermatoses. Cases with occupational dermatosis were further evaluated regarding using a PPE kit, and DLQI was calculated.

**Results:** 19% of HCWs had dermatoses associated with PPE and hygiene practices. Hands were most affected, followed by the face, nasal bridge, and facial skin in contact with goggles. 48% had Mathias score >/= 4. Most cases had reported some impact on their quality of life. A significant association could be established between frequency of hand washing >/= 10 times/day with hand dermatitis (p=0.000).

**Conclusion:** The use of PPE has significantly raised cases of occupational dermatosis among HCWs. Repeated hand washing and hand sanitizer use has increased the incidence of hand dermatitis.

## Introduction

The current coronavirus disease 2019 (COVID□19) has caused significant changes in practices and preventive strategies among Health care Workers (HCWs). It has been demonstrated that these practices have resulted in an increase in various occupational dermatoses in them. Skin problems often significantly impact daily activities, potentially limiting the effective workforce and efficiency. HCWs caring for COVID□19 patients must wear, for many hours, specific PPE (PPE) daily and are therefore susceptible to PPE□related adverse skin reactions.(1,2,3,4,5) Hand hygiene with alcohol□based hand rubs and frequent handwashing with water and soap also lead to hand dermatitis varying from relatively mild to debilitating. So, in this study, we intended to look at the prevalence of occupational contact dermatoses among HCWs in our tertiary care institute, which was recently designated as one of the Covid-dedicated hospitals. There are studies worldwide, but few from India looked into this matter. This aspect must be addressed during COVID management and future healthcare practices.

## Materials And Methods

This single-center cross-sectional study screened 1105 HCWs of tertiary care hospitals in Central India. All HCWs (doctors, nursing staff, sanitation workers) who had worked on COVID duty, are currently in quarantine, or resumed regular duty have been included. The study continued for two years after the Institutional Human Ethical Committee’s approval (Ethics approval number LOP/2020/IM0299). HCWs were asked to fill out a predetermined questionnaire either in Google form (for those in quarantine) or hard copy (who had resumed regular duty). The persons who experienced dermatological complaints were noted and examined or asked to share clear images of their lesions. Other information like duration, type, hours of duty, types of PPE use, frequency of hand washing, type of hand sanitizer/soap used, personal or family history of atopy, and previous history of dermatitis, if any were recorded. Mathias criteria made the final diagnosis of occupational contact dermatitis, and the impact of the occupational dermatosis on quality of life was calculated using Dermatology Life Quality Index. The diagnosis of these dermatoses was mostly made clinically by an experienced dermatologist. When allergic contact dermatitis is suspected, Patch testing with Indian standard series and suspected material was done. Statistical analysis was performed using SPSS version 25. Descriptive statistics were expressed as frequency and percentages. An Association study between the categorical variables was performed. Statistical significance was calculated using the chi-square test and Fisher’s exact test, and a p-value of <0.05 was taken as statistically significant.

## Results

Among the 1105 HCWs screened, 212 (19.20%) had dermatoses associated with PPE and hygiene practices. Background details of the screened population are described in Table 1.

**Table 1:**
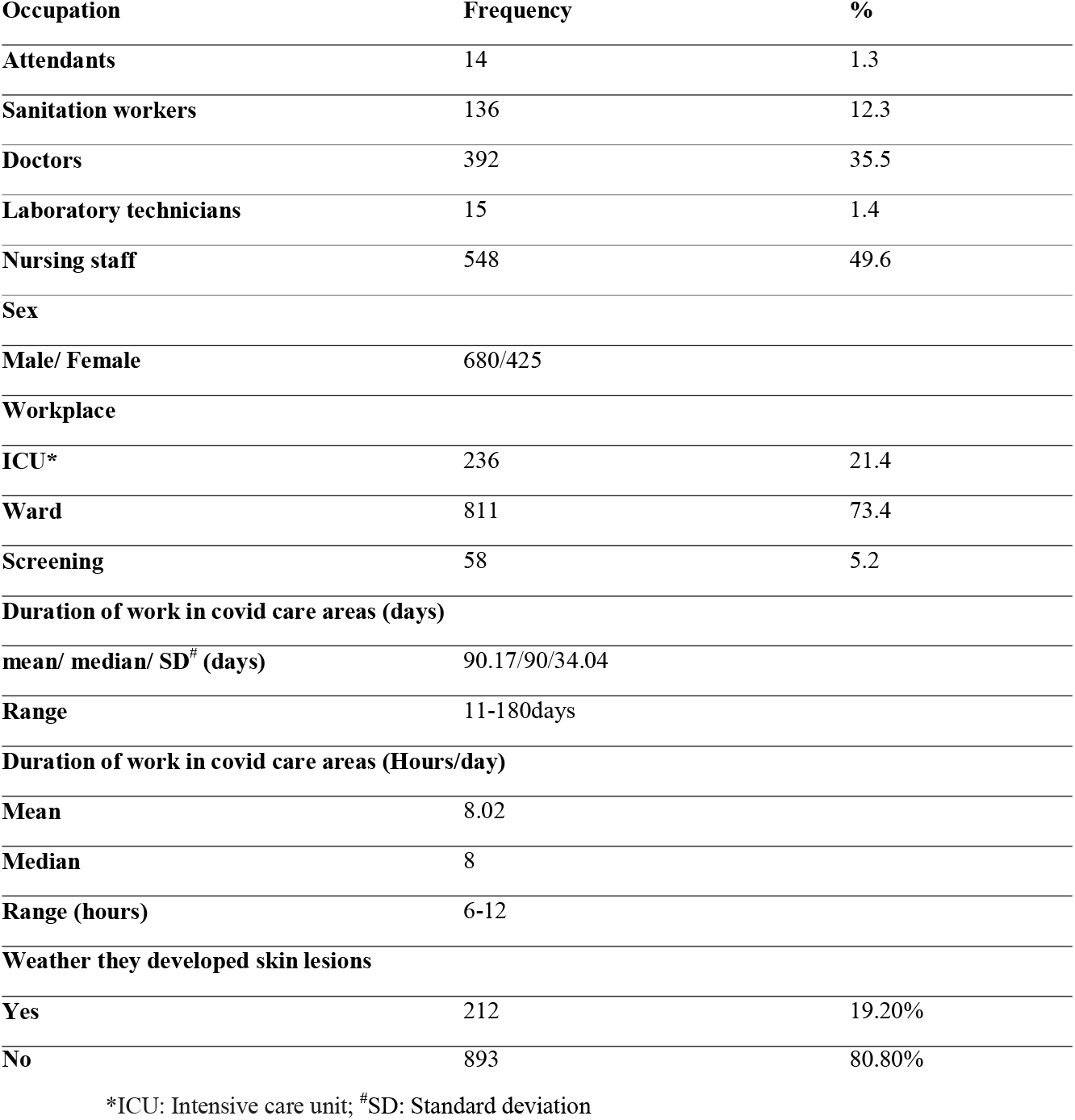
Demographic description of the HCWs associated with occupational dermatosis.

Among HCWs developed dermatoses (212), the hands were affected in 102, the face in 67, the nasal bridge in 43, and facial skin in contact with the goggle margin in 19 cases (Figure 1).

**Figure 1:**
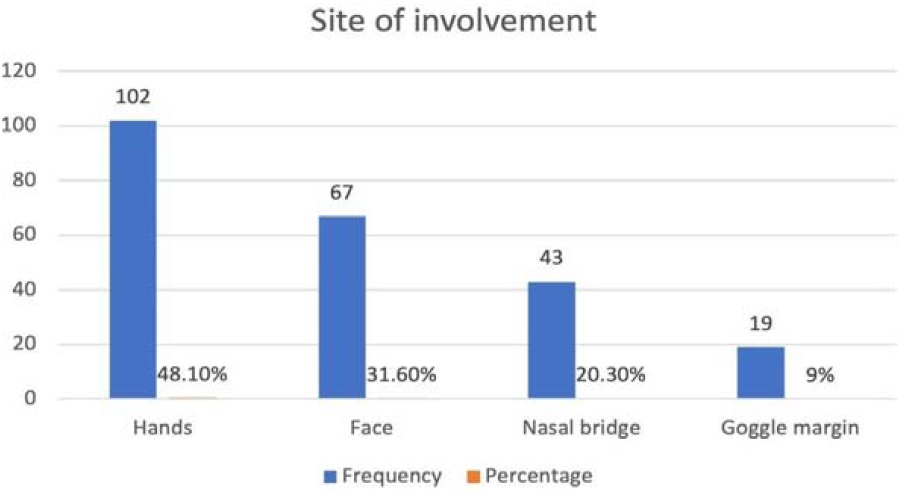
Site of Involvement. Hands were involved in maximum participants followed by the face.

Over hands, the most common site was web spaces (69 cases) followed by fingers in 54 and the dorsum of the hand in 47 cases (Table 2).

**Table 2:** Distribution and types of skin lesions in healthcare workers.

| Site of Involvement |  | Frequency | Percentage |
| --- | --- | --- | --- |
|  | Hands | 102 | 48.10% |
|  | Face | 67 | 31.60% |
|  | Nasal bridge | 43 | 20.30% |
|  | Goggle margin | 19 | 9% |
| Symptoms | Scaling | 61 | 28.80% |
|  | Papules | 62 | 29.20% |
|  | Pain | 110 | 51.90% |
|  | Burning | 78 | 36.80% |
|  | Erythema | 96 | 45.40% |
|  | Fissuring | 7 | 3.30% |
|  | Itching | 56 | 26.40% |
|  | Hyperpigmentation | 1 | 0.50% |
|  | Vesicles | 14 | 6.60% |
|  | Thickening | 2 | 0.90% |
|  | Erosions | 2 | 0.90% |
|  | Excoriations | 1 | 0.50% |
|  | Edema | 1 | 0.50% |
|  | Dryness | 1 | 0.50% |
| History | Similar illnesses in past | 46 | 21.7 |
|  | Hand dermatitis | 16 | 7.5 |
|  | Atopy | 71 | 33.5 |
|  | Family history of Atopy | 49 | 23.1 |
| Frequency of hand washing | Mean | 12.89 |  |
|  | Median | 12 |  |
|  | SD* | 4.2 |  |
|  | Range | 5-30times/day |  |
| Frequency of hand washing category | <10 times/day | 80 cases |  |
|  | >= 10 times/day | 132 cases |  |
| Material used of hand washing | Soap | 212 |  |
|  | Sanitizer | 212 |  |
| Site of Involvement |  |  |  |
| Hand (102) | Palm | 26 | 12.3 |
|  | Fingers | 54 | 25.5 |
|  | Web spaces | 69 | 32.5 |
|  | Dorsum of hand | 47 | 22.2 |
| Face (129) | Nose | 77 | 36.3 |
|  | Cheeks | 31 | 14.6 |
|  | Forehead | 21 | 9.9 |
| Examination | Papules | 89 | 42 |
|  | Erythema | 159 | 75 |
|  | Scaling | 70 | 33 |
|  | Lichenification | 2 | 0.9 |
|  | Vesicles | 15 | 7.1 |
|  | Fissuring | 13 | 6.1 |

\*SD: standard deviation

Over the face, the nose was more commonly involved, followed by the cheeks and forehead. The most commonly reported symptoms were pain (110), redness (96), burning (78), and tiny eruptions (62), whereas, on examination, erythema was the most typical finding (159 cases), papules in 89, and scaling was evident in 70 cases (Figure 2).

**Figure 2:**
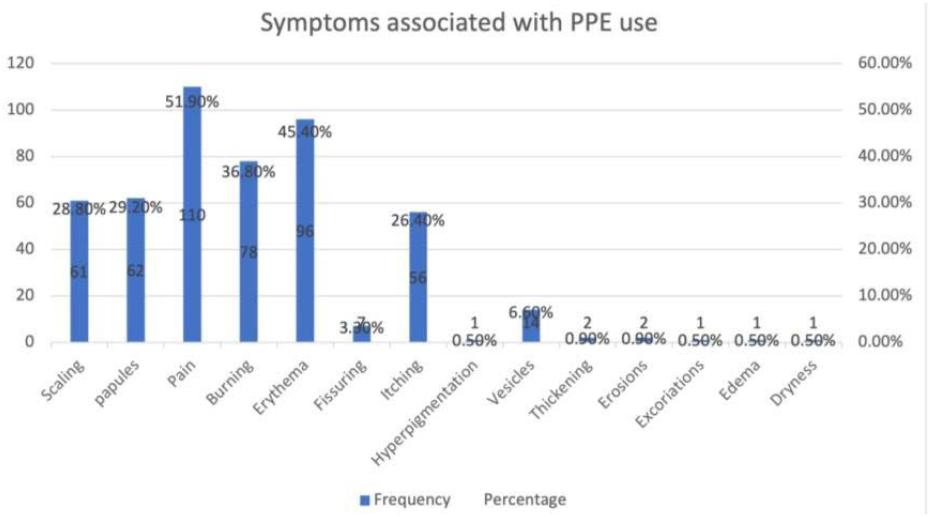
Frequency of various symptoms associated with PPE use. Pain and erythema were reported the most, while excoriations, edema, and dryness were reported the least. PPE: personal protective equipment

As per Mathias criteria (>/= 4 scores), only 48% of cases had dermatosis qualified to be occupation-related (Table 3).

**Table 3:** Mathias criteria among the affected healthcare workers.

|  |  |  |
| --- | --- | --- |
| **Mathias score** | Mean | 2.62 |
|  | median | 2 |
|  | Standard deviation | 1.377 |
|  | Range | 1-4.0 |
| **Categories** | <4 | 110(51.88%) |
|  | >4 | 102(48.11%) |

We also assessed the impact of dermatoses on the quality of life of HCWs. A significant proportion of cases (70%) had a small impact on quality of life (DLQI score 2-5), and a small group of cases had a moderate and tremendous impact on quality of life, 20 and 1 %, respectively. As per the diagnosis, hand dermatitis (48.10%) was the most prevalent, followed by pressure-induced erythema in 34.40% and mask-induced acne in 31.60% (Figure 3).

**Figure 3:**
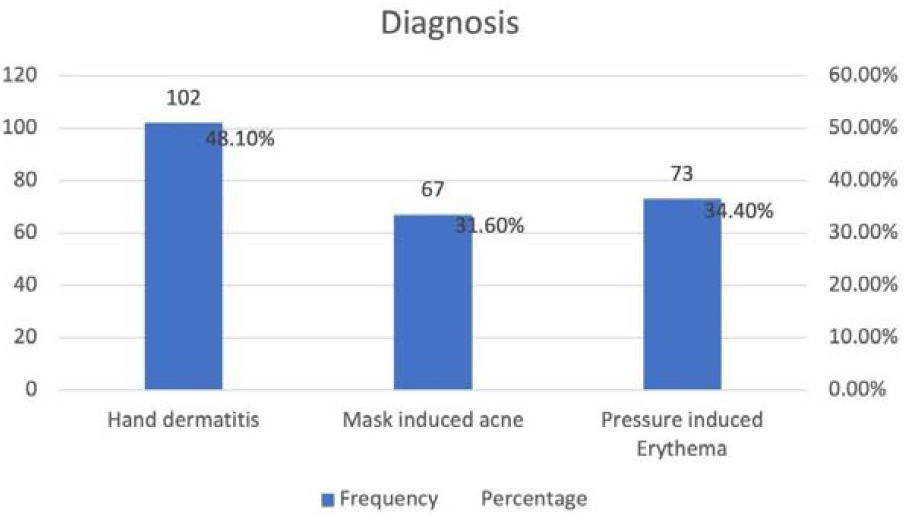
Frequency of Various dermatoses reported due to PPE use. Maximum healthcare workers were found to have hand dermatitis. PPE: personal protective equipment

Among those with hand dermatitis, the mean frequency of hand washing was 12.89, with 62.2% having >10 times per day frequency of hand washing (Figure 4).

**Figure 4:**
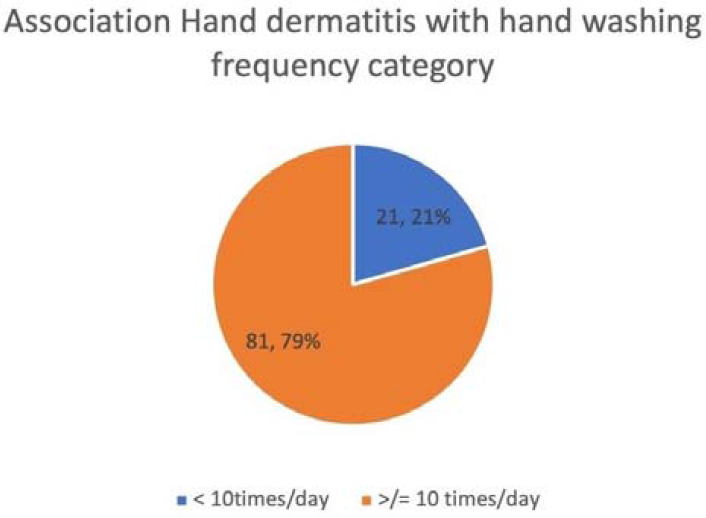
Frequency of hand washing among cases with hand dermatitis. A large proportion of healthcare workers with hand dermatitis were found to have washed their hands more than ten times a day.

A significant association could be established between the frequency of hand washing >/= 10 times per day with hand dermatitis(p=0.000). The incidence of dermatosis in various occupation groups hand dermatitis and pressure-induced erythema were common among nurses (p=0.000, 0.025 respectively) as compared to other groups of HCWs, whereas mask-induced acne was more common among doctors (p=0.000) (Figure 5).

**Figure 5:**
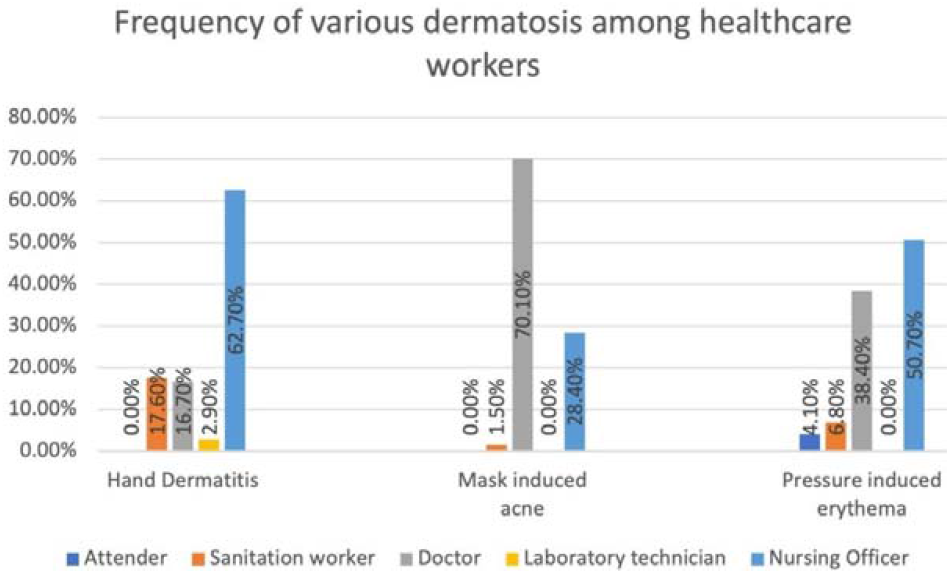
Frequency of Dermatosis among various cadres of healthcare workers. Nursing officers and doctors were found to have all the three dermatoses

Most of the cases with dermatoses were from the ward, followed by ICU (Figure 6).

**Figure 6:**
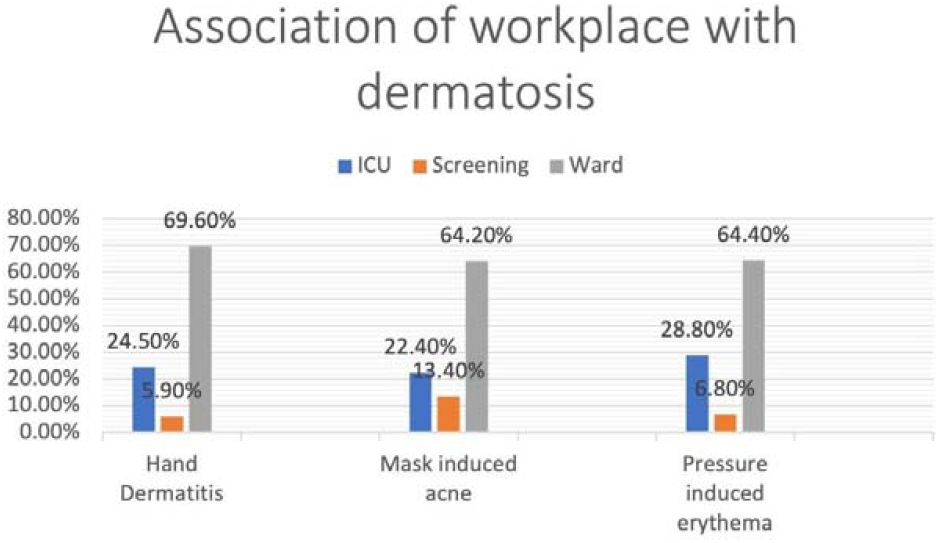
Association of the workplace with dermatosis. Healthcare workers working in wards were maximally found to have dermatoses due to personal protective equipment. Dermatoses followed this in ICU healthcare workers. ICU: Intensive care unit.

Figures 7, 8 and 9 show cases of mask-induced acne, pressure-induced erythema over the back, and hand dermatitis post latex gloves use.

**Figure 7:**
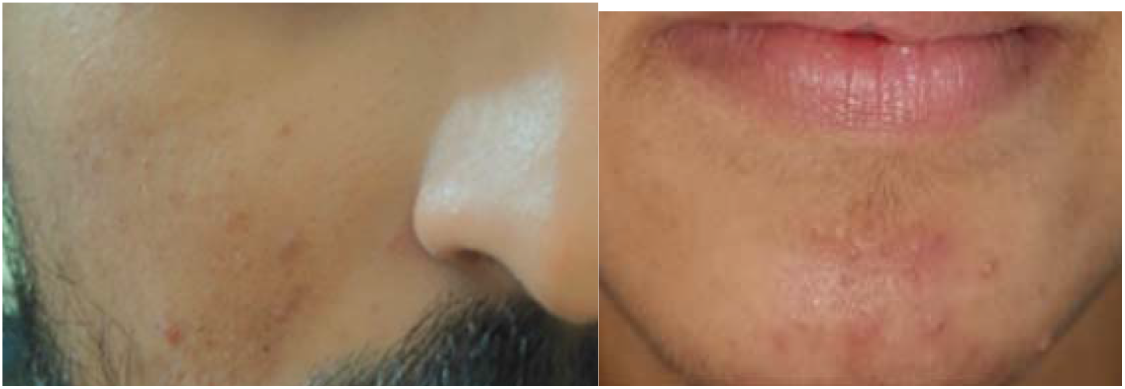
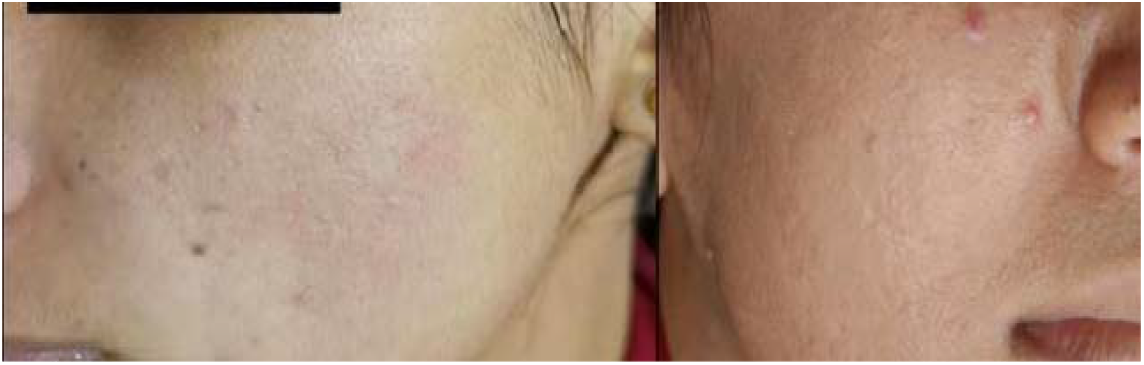
Cases with mask-induced acne. Acne exacerbation along the areas of mask vis-à-vis cheeks.

**Figure 8:**
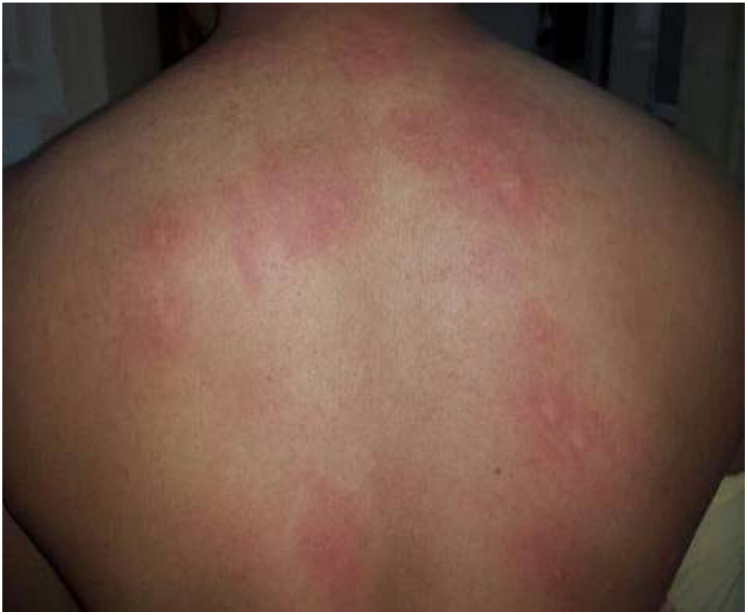
Cases with pressure-induced erythema over the back. Erythema over nasal bridge after using face mask. Few urticarial plaques and erythema also present over back after using personal protective equipment.

**Figure 9:**
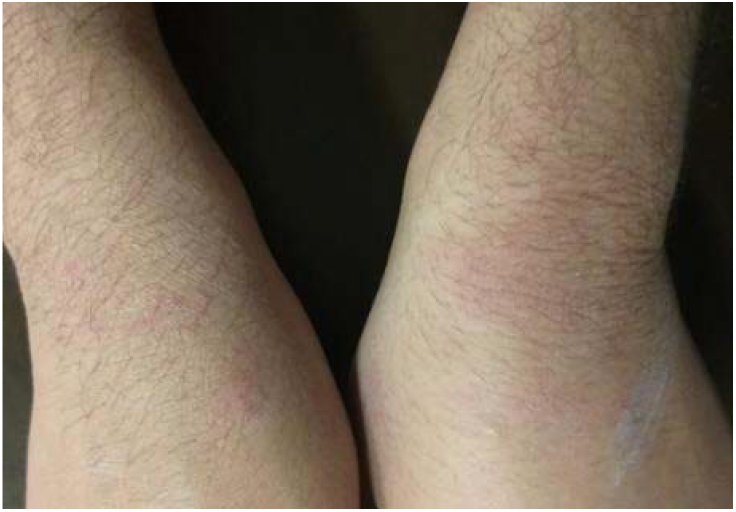
Case with hand dermatitis post latex gloves use. Ill-defined erythema with papules and fine scaling present over the dorsum of hands after using powdered latex gloves.

## Discussion

Though the pandemic is slowly weaning off, the experience regarding occupational dermatoses in HCWs would help if future healthcare planning. We performed a cross-sectional observational study among covid HCWs of a tertiary care center in Central India, where we screened 1105 HCWs for PPE-associated dermatoses and found 212 such cases (19.15%). In various studies, 73.5% to 97% of health professionals experienced dermatoses. (1,2,3,4,6) The current study’s lower prevalence of hand dermatitis could be because of increased awareness of skin care and preventive measures like using barrier creams and oils, using pads under goggles, etc, by HCWs. Lan et al. reported most common skin problems were dryness and peeling over the nasal bridge, hands, cheeks, nose bridge, and forehead. (3) Atzori et al. reported dryness, burning, and itching as the most identified symptoms among HCWs, and frequently affected areas were the bridge of the nose, cheeks, hands perioral, and periocular regions. (7) Most frequently reported symptoms in our series were pain, erythema, burning, and papules in decreasing order of frequency. In another study, the problematic areas of occupational dermatosis were commonly over the face, followed by the hands, legs, and trunk. (2) We found that hands were almost commonly affected areas, followed by cheeks and nasal bridges. On examination, erythema was found in a maximum number of patients (159 cases), followed by papules (89) and scaling (70). 48% of cases had dermatosis qualified to be occupation related as per Mathias criteria (>/= 4 scores). Mask-associated dermatoses were detected over the nose, cheeks, retro-auricular region, over nasal bridge among those wearing glasses and over the forehead in visor wearers. (1) Foo et al. reported that nearly 1/3rd of HCWs using N95 masks developed acne, dermatitis, or pigmentation, with acne reported to be the most common dermatoses. Friction and occlusion were suspected as the underlying cause. (8) Singh et al. reported contact dermatitis (39.5%) as the most common manifestation and followed by friction (25.5%) dermatitis. The nasal bridge (63%) was reported as the commonest location among the nose. (9) O’Neill et al. reported 17% HCWs with acne and 3% with facial pressure injuries due to PPE worn during the covid-19 pandemic. (10) Nose was also the most common site associated with mask-related dermatosis. Due to occlusion, the mask provides a moist and warm environment that traps bacteria and sebum, worsening or triggering symptoms for contact dermatitis, mask-induced acne, and pressure injuries due to tight masks. (10,11) Damage to the epidermis was more prevalent among HCWs wearing N95 masks and goggles for more than 6 hours/day. Face masks and headgear were worn tightly for multiple hours, resulting in pressure urticaria, friction dermatitis, irritant, allergic contact dermatitis, abrasions, and aggravation of pre-existing dermatoses. (12,13) Increased cases of hand dermatitis in all the published studies are most likely due to increased hand hygiene practices, duration of gloves use, and wet work. (14) We looked into the detail of the hand hygiene practices among affected individuals. The mean frequency of hand washing in our series was 12.89, with 62.2% having>10 times /day of hand washing. A significant association could be established between frequency of hand washing >/= 10 times per day with hand dermatitis(p=0.000).

Previously also, significant risks associated with hand washing more than ten times/day have been reported. (7,12,15) Repeated use of alcoholic agents induces stratum corneum proteins denaturation, impaired keratinocyte maturation, and changes in intercellular lipid composition, which leads to barrier dysfunction and release of inflammatory cytokines leading to hand dermatitis. (16) Though gloves act as protective barriers, the prolonged wearing of gloves can also disrupt the barrier; even the gloves can also add to the deterioration of the barrier when the skin becomes irritated and increases susceptibility to allergens and other irritants. (10) Irritant contact dermatitis was detected in 98% of the employees in a study conducted in HCWs and was associated with frequent hand washing. (17,18) Moisturizers containing humectants, oils, and fats can effectively replace depleted skin lipids and maintain skin hydration, which helps in restoring the skin barrier. (16,19) These dermatoses could be troublesome and difficult to cope with for healthcare workers. Munise et al. reported 73% of cases with small or no effect on the quality of life based on the DLQI score. (1) In our study, 70% of cases had a small impact on quality of life, similar to the previous reports.

### Limitations

The inability to do patch tests in all suspected individuals due to quarantine and other covid related restrictions was the limitation of our study. All the lesions could not be examined in person. Sometimes, when they could be examined, the severity was reduced.

## Conclusion

This is likely the foremost study from the Indian subcontinent on occupational dermatosis in covid health care workers, which focused on etiology, manifestations, change in practice pattern, and its impact on the quality of life. Though the covid pandemic is settling off gradually, learning from this aspect could help in better management in the future. Educating HCWs regarding proper skin care management may effectively prevent occupational skin disorders. Additional skin moisturizing may be needed for HCWs at risk of hand dermatitis to hygiene products.

## Data Availability

All data produced in the present study are available upon reasonable request to the authors

